# Disparities in cancer genomics by ancestry in the 100,000 Genomes Project

**DOI:** 10.1101/2024.11.14.24317310

**Authors:** T. Nguyen, Sam Tallman, Yoonsu Cho, Alona Sosinsky, John Ambrose, Steve Thorn, Maxine Mackintosh, Matthew A. Brown, Loukas Moutsianas, Matt J. Silver, Karoline Kuchenbaecker

**Affiliations:** Genomics England, UK; Medical Research Council Integrative Epidemiology Unit, University of Bristol, Bristol, UK; Department of Oncology, University of Oxford, UK; Alan Turing Institute, UK; Department of Population Health, London School of Hygiene & Tropical Medicine, UK; Medical Research Council Unit The Gambia at the London School of Hygiene and Tropical Medicine, The Gambia; Division of Psychiatry, University College London, UK; UCL Genetics Institute, University College London, UK

## Abstract

**Purpose:** Most research on genetic screening and precision oncology is based on participants of European ancestry, making it vital to evaluate the performance of these approaches in diverse populations. We analysed data from the 100,000 Genomes Project (100kGP) to assess ancestry-related differences in cancer variant prioritisation.

**Patients and Methods:** To assess the representativeness of the 14,775 participants with cancer from the 100kGP, we compared recruitment ratios for self-reported ethnicities to those in England. For genetic ancestry groups we analysed differences in detection rates for potential pathogenic variants (PVs) in the germline and somatic mutations in genes with treatment implications and investigated possible causes of observed disparities.

**Results:** Recruitment rates for Black and Asian ethnicities compared with White ethnicity in the 100kGP were consistent with rates in England, except for bladder and prostate (Black and Asian) and breast (Asian only) where Black and Asian ethnicities were recruited at higher rates than expected compared to White ethnicity.

Patients with non-European genetic ancestry were more likely to carry variants classified as potential pathogenic compared to European ancestry (p=0.006). PVs were identified in 4.6% of South Asian (adjusted model: odds ratio=1.88, 95%CI=1.21-2.93) and 5.3% of African ancestry patients (odds ratio=2.24, 95%CI=1.44-3.48) compared with 2.2% in European.

Fewer non-synonymous somatic mutations in actionable genes were identified in patients of non-European ancestry (p=0.004). WGS failed to identify treatment-relevant findings for 26% of patients of South Asian ancestry compared with 16% of European ancestry.

**Conclusion:** The excess germline variants classified as PVs in patients with non-European ancestry may impede the diagnostic process. Our analysis demonstrates the need for better variant classification across diverse ancestries to ensure equitable implementation of genomics in cancer care.

## Introduction

Identification of carriers of cancer susceptibility variants through genome sequencing facilitates prevention and earlier disease diagnosis. Characterising somatic mutations in tumour tissue plays an important role in targeted treatment. England was the first country to offer whole-genome sequencing (WGS) as part of routine cancer care within a national health care system, through the English National Health Service’s Genomic Medicine Service (GMS)^1^.The foundation for this was laid by the 100,000 Genomes Project (100kGP)^2^, which recruited more than 15,000 patients to the 100kGP Cancer Programme between 2013-2018^3^14/11/2024 03:04:00. Several other countries are now following suit to increase the use of genome sequencing in oncology^4^.

Previous studies have identified differences between ethnicities in terms of cancer incidences, diagnosis and prognosis^5^ ^6, 7^ ^8^. It is vital that genetic carrier screening and precision oncology provide equal benefits for people from diverse ethnic backgrounds to avoid exacerbating existing health inequalities. USA-based studies found that carrier screening using gene-panels yields variants of uncertain significance (VUS) about twice as often for Black and Asian compared to White individuals^9^. Non-White patients also had slightly lower rates of likely pathogenic variants (PVs)^10^. However, a more recent study did not find a statistically significant difference in PVs between Black and White women with breast cancer^11^.

The role of ancestry for carrier screening and precision oncology has never been investigated for a national cancer sequencing programme. Furthermore, the causes of any differences remain largely unknown, so that routes towards improving performance for diverse ethnic and ancestral groups are unclear.

Here, we examine how representative the 100kGP is in terms of self-reported ethnicity. We further assess whether there are ancestry differences in the identification of clinically relevant germline variants in high-risk genes, such as *BRCA1/2*, as well as somatic cancer variants and investigate how these relate to demographics, tumour characteristics, population genetic differences and features of the bioinformatics pipeline used for variant prioritisation.

## Methods

### Cohort

We analysed WGS data from cancer patients recruited into the 100kGP which have been described in detail elsewhere^3^ (Supplementary Table 1). Our analysis was limited to patients in the version 15 release of the National Genomic Research Library (NGRL)^12^, a database of WGS and linked health data from consenting National Health Service (NHS) patients. Each patient had one germline and one or more tumour samples which were sequenced at 30× coverage and 100×, respectively. Cancer types were assigned according to the location of the primary tumour. Basic patient and sample data were collected at the time of DNA sample submission. Secondary clinical information, including stage and grade, was gathered from the NHS England and Public Health England National Cancer Registration and Analysis Service (NCRAS)^3^.

Twelve patients with conflicting sex and cancer types or with multiple cancer types were excluded. Patients were assigned female or male sex if their self-reported sex at registration matched their sequenced sex karyotype. Patients were assigned indeterminate sex if they did not report a sex, their sequenced karyotype did not match their self-reported sex, or their sequenced sex karyotype was not XX (Female) or XY (Male).

### Ethnicity representation analysis

100kGP participant ethnicity was self-reported at registration using UK Office of National Statistics (ONS) categories^13, 14^. Recruitment rate ratios of Black to White and Asian to White ethnicities were compared to ratios in England during 2013-2017 reported by Delon *et al*. ^5^. The age-adjusted recruitment rate ratio of ethnicity *e* to White ethnicity within the 100kGP for the target sex and cancer type were calculated as

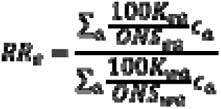

with

- *100KG_ea_*: total cancer cases within the 100kGP for ethnicity *e*, age group *a*, and target sex and cancer type
- *ONS_ea_:* ONS^15^ estimate of the English population size for ethnicity *e*, age group *a*, and target sex, averaged across years 2013-2018
- *C_a_*: weight of age group *a* (5-year interval) within the 2013 European Standard Population^16^, *C_a_∈ [0, 1],*
- *100KG_wa_*: total cancer cases within the 100kGP for white ethnicity, age group *a*, and target sex and cancer type
- *ONS_wa_*: ONS estimate of the English population size for white ethnicity, age group *a*, and target sex, averaged across years 2013-2018

Only cancer types with at least 100 patients and with matching ICD10 codes were compared. We followed the same methods used by Delon *et al.* to adjust for age and to calculate confidence intervals (Supplementary Methods). The difference between log-rate ratios from the 100kGP and England were compared using z-tests^17^. We applied a Bonferroni correction to P-values to account for multiple testing.

### Ancestry assignment

Individuals were assigned to genetically-inferred ancestry groups based on their similarity to super-populations from the 1000 Genomes Project as previously described^18^ (Supplementary Table 2). Any participant not meeting the threshold for similarity to a single reference population was classified as ‘unassigned’. For brevity, we occasionally use ‘patients with European ancestry’ etc to refer to patients assigned to the European ancestry group, while noting that no one derives from a single ancestry in any meaningful sense. When referring to ancestry in this manuscript, this always implies genetically-inferred ancestry.

### Variant prioritisation and variant counts

Germline and somatic variants were identified and classified by the 100kGP cancer bioinformatics pipeline (v1.6–v1.11), as previously described^19^. Only the latest pipeline version outputs were analysed for each sample. Functional, allele frequency, and variant database annotations were obtained from the NGRL^20^.

Germline variants were classified as likely pathogenic/pathogenic variants or “tier 1” if they were found in genes linked to the patient’s cancer type according to PanelApp, a gene panel database for health disorders^21^, and were either predicted protein truncating variants for which the mechanism of pathogenicity is loss of function (excluding variants annotated as benign or likely benign in Clinvar^22^ with a rating of at least two stars) or were listed in ClinVar as pathogenic or likely pathogenic (with a rating of at least two stars) (Supplementary Figures 1 and 2). We refer to these as germline potential pathogenic variants (PVs). Candidate or “tier 3” variants are a lower priority classification, indicating variants in genes within a broader cancer susceptibility panel, across a wider range of consequence types, where the frequency in an internal Genomics England dataset of >6,000 unrelated individuals is <0.05% for dominant-acting genes and <2% for recessive genes. Variants listed in ClinVar as benign or likely benign with a rating of at least two stars were excluded from this category.

Somatic mutations were classified as actionable cancer-related somatic variants, or “domain 1”, if they were protein-altering and located in genes affecting diagnosis, prognosis, or treatment for the patient’s cancer type or lead to eligibility for a clinical trial (Supplementary Figure 3). Cancer-related somatic “domain 2” mutations were protein-altering mutations located in genes implicated in any cancer type. “Domain 3” mutations were protein-altering mutations found in any protein-coding genes.

### Statistical analyses of cancer variants

In the analyses for variant prioritisation, patients with cancer types with fewer than 5 individuals, haematological cancers (n=788), childhood cancers (n=154), unknown primary carcinomas (n=84), patients with indeterminate sex (n=77), and patients missing somatic mutations in genes were excluded. Patients of admixed Latin American ancestry (n=35) were also excluded due to low sample numbers. The association of genetic ancestry with the probability of finding at least one PV was modelled by logistic regression with cancer type as a random effect. Negative binomial regression was used to evaluate the association between ancestry and the total number of prioritised variants per patient (candidate germline variants, non-synonymous somatic variants in actionable genes, and all domains). For all models, ancestry was represented as a categorical predictor with the largest group, European ancestry, as a reference.

Likelihood ratio tests (LRTs) were used to test for an overall effect of ancestry on outcomes by comparing baseline models adjusted for sex and cancer type to the same model with an additional covariate for ancestry. For significant LRTs (p<0.05) we assessed differences between individual groups using the regression coefficients for ancestry.

We explored whether ancestry differences in germline variants are linked to other variables by adding them as covariates to the model: age at registration, genetic diversity (ratio of heterozygous/homozygous variants), and total number of germline variants. For somatic mutations, we considered: age at sampling, tumour mutation burden, tumour grade, and cancer type. Tumour mutation burden (TMB) was defined as the count of somatic mutations in any region of the genome that were smaller than 50bp and passed variant calling filters.

An additional sensitivity analysis was conducted in female breast cancer patients to assess whether ancestry group effects on PV detection were confounded by age. For this we created early (age ≤46 years, N=394) and late (age ≥53 years, N=2020) onset groups and repeated the analysis described above.

We estimated population allele frequencies for identified candidate germline variants using data from the Genome Aggregation Database (gnomAD) v3.1^24^. 100kGP ancestry groups were mapped to gnomAD super-populations with the same names, except for the European ancestry group which was mapped to ‘Non-Finnish European’ in gnomAD.

### Code availability

Code for all analyses is available at https://gitlab.com/genomicsengland/Data_Diversity_Public/cancer-100kg-diversity-blog/-/tree/2024paper?ref_type=heads

### Data availability

The data supporting the findings of this study are available within the Genomics England Research Environment, a secure cloud workspace. Details on how to access data for this publication can be found at https://re-docs.genomicsengland.co.uk/pan_cancer_pub/. To access the genomic and clinical data within this Research Environment, researchers must first apply to become a member of either the Genomics England Research Network (previously known as the Genomics England Clinical Interpretation Partnership, GECIP) (www.genomicsengland.co.uk/research/academic) or a Discovery Forum industry partner (www.genomicsengland.co.uk/research/research-environment). The process for joining the Genomics England Research Network is described at www.genomicsengland.co.uk/research/academic/join-gecip. Data that have been made available to registered users include: alignments in BAM or CRAM format; annotated variant calls in VCF format; signature assignment; tumour mutational burden; sequencing quality metrics; summary of findings shared with the Genomic Lab Hubs; and secondary clinical data as described in this paper.

## Results

### Ethnicity representation

We compared the ethnic composition of 14,775 cancer patients in the 100kGP (Figure 1) to the ethnic composition of cancer patients in England published by Public Health England (PHE)^5^. There was no evidence that diverse ethnic groups were underrepresented when compared with the national statistics (Figure 2, Supplementary Table 3). In fact, the recruitment rate ratio for breast cancer was 2.2 for Black vs White women in 100kGP compared with 0.81 for Black vs White women in the PHE data (fold-change in rate ratios=2.7, p=6×10^-10^), suggesting higher representation of Black women in 100kGP than expected given the ethnicity-specific incidence rates in England. Compared with PHE, the 100kGP also had higher recruitment rates of Black vs White men with prostate cancer, Black vs White men with bladder cancer, Black vs White women with breast cancer, and Asian vs White women with breast cancer (fold-change=3.7, p = 0.012; fold-change=6.1, p=0.047; fold-change=1.4, p=0.024, respectively).

**Figure 1.**
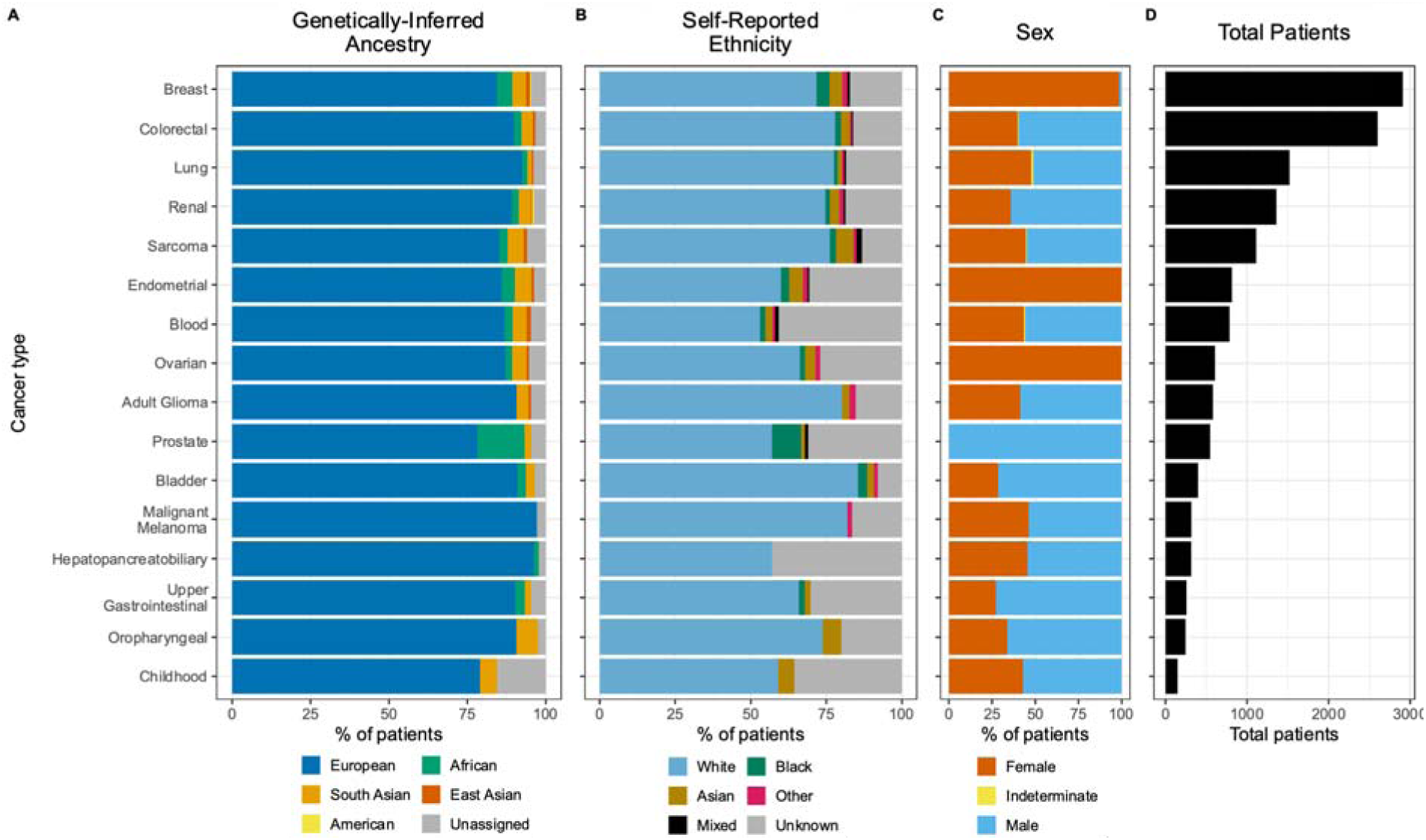
Patient characteristics by cancer type. **A**. Percentage of patients assigned to each genetically-inferred ancestral super-population group in the 100kGP Cancer Programme. **B**. Percentage of patients by self-reported ethnicity **C**. Percentage of patients by sex. **D**. Total number of patients. Cancers with fewer than 100 patients not displayed. Sub-groupings with fewer than 5 patients for a cancer type are not included in percentage calculations. Tabular data in Supplementary Tables 1-2

**Figure 2:**
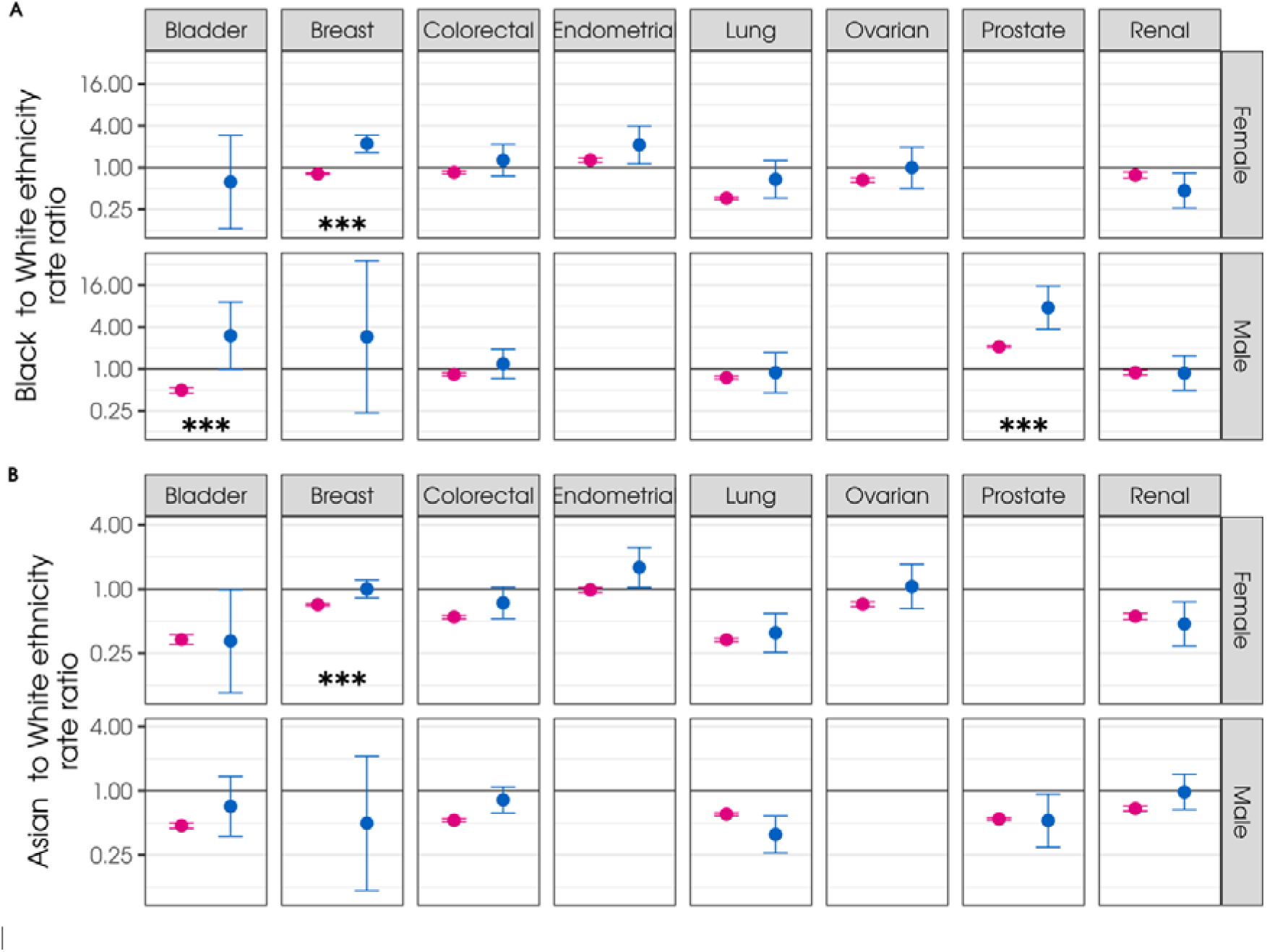
Recruitment ratios for self-reported A) Black vs White and B) Asian vs White ethnicities. Blue points show age-adjusted recruitment ratios for the 100,000 Genomes Project (100kGP); pink points indicate age-adjusted incidence ratios for England from Public Health England (PHE)^5^. Error bars show 95% confidence intervals. Comparisons for sub-categories with fewer than 5 patients in the 100kGP are omitted. *** indicates a statistically significant difference between PHE and 100kGP ratios at alpha=0.05 after correction for multiple tests across cancer types.

### Ancestry differences in germline variant prioritisation

Overall, 2.4% of 13,645 patients included in this analysis had one or more PV. 70.8% of patients had at least one candidate germline variant (Supplementary Table 4). The most common genes with PVs were *BRCA2* in European (0.5%) and African (1.8%) and *BRCA1* in South Asian ancestries (1.2%). The most common genes with candidate variants were *ATM* in European (7.5%), South Asian (7.6%) and East Asian (10%) ancestries and *PALB2* in African ancestry (12%).

Ancestry had a significant association with the likelihood of carrying a variant classified as PV (likelihood ratio test (LRT) p=0.002). African and South Asian ancestries had significantly higher frequencies of PVs compared to European ancestries (Figure 3A, Supplementary Table 5; odds ratio (OR)=2.24, p=3×10^-4^ and OR=1.88, p=5×10^-3^, respectively). PVs were identified in 4.6% of South Asian and 5.3% of African ancestry patients compared with 2.2% in European. Furthermore, all of the ancestry groups had significantly more candidate variants compared to the European ancestry group (p<3×10^-12^, Figure 3B, Supplementary Table 6).

**Figure 3.**
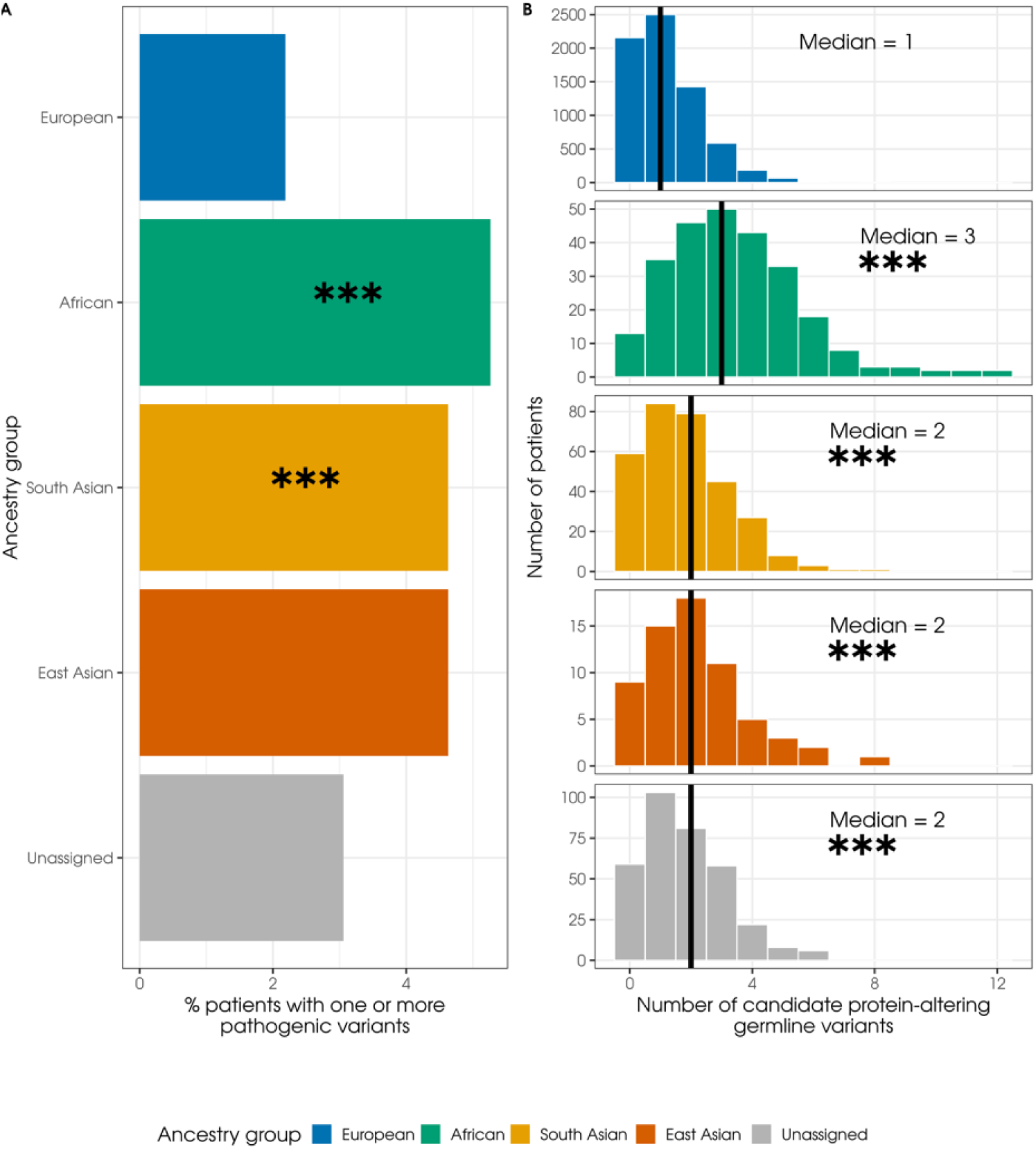
Percentage of potential pathogenic variants (PVs) and number of candidate variants by ancestry group. **A:** Percentage of patients with at least one PV by ancestry group. **B:** Distribution of prioritised candidate germline variants by ancestry group. The median number of variants is marked with a vertical black line. *** indicates a significant difference between the number of variants for a given ancestry compared to individuals of European ancestry (model accounting for sex and cancer type). Tabular data in Supplementary Tables 4-6.

We assessed whether these differences might be linked to greater genetic diversity or different segregation models in some ancestry groups^25^. However, in models adjusted for the total number of germline variants and for the heterozygous/homozygous ratio, non-European ancestry groups consistently had more PVs and candidate variants than those in European ancestry groups and neither covariate was associated with the likelihood of having a PV (Supplementary Tables 5 and 6).

We next investigated the effect of age on the likelihood of finding PVs in breast cancer patients, the largest cancer type in the 100kGP. The most prevalent PVs were in the *BRCA1* and *BRCA2* genes which are linked to earlier age at diagnosis^26^. The likelihood of a PV was associated with younger age across all cancers (p=2×10^-17^). To separate the effect of age from ancestry, female breast cancer patients were split into early and late onset groups. South Asian and African ancestries were more likely to have PVs than European ancestry in both age groups, but only South Asian ancestry in the early onset group and African ancestry in the later onset group remained statistically significant in this small data set (Supplementary Table 7). For the East Asian ancestry group there were only 12 and 17 participants, respectively, in each of these age subgroups and none of them carried PVs.

We next calculated the allele frequencies (AFs) of candidate germline variants (Figure 3B). High-risk variants in cancer predisposition genes are expected to be rare. Contrary to this, we found that each non-European ancestry group had a proportion of candidate variants with AFs greater than the 2% common variant threshold used by the variant prioritisation pipeline. Most notably, 25% of candidate variants in patients of African ancestry were common in the African gnomAD group (Figure 4, Supplementary Table 8). This can be explained by the fact that reference data used by the prioritisation pipeline for identifying common variants is not separated by ancestry and is predominantly based on European ancestry samples.

**Figure 4.**
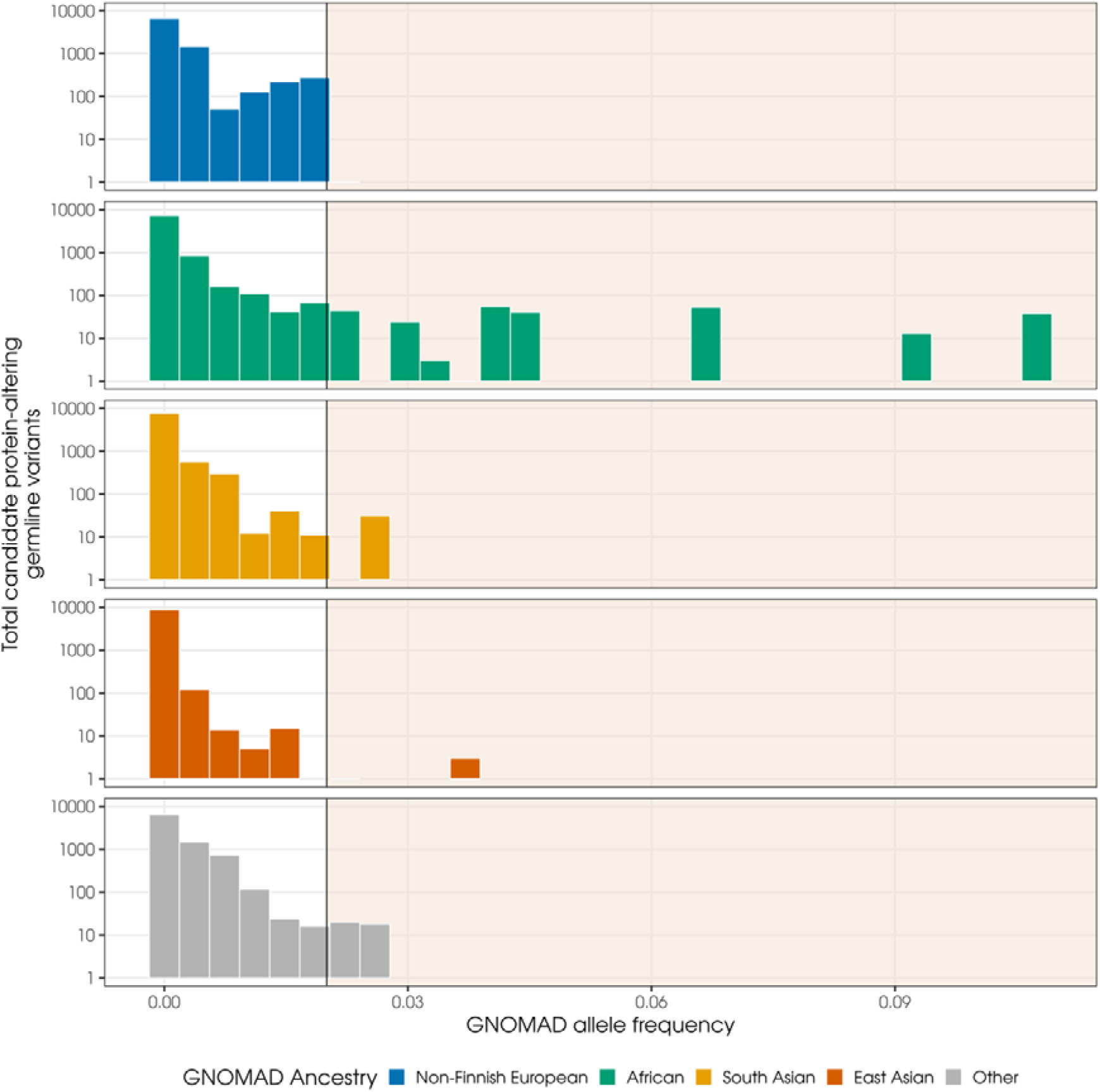
Distribution of ancestry-specific allele frequencies in gnomAD for candidate germline variants, stratified by ancestry group. The shaded region covers variants which are more common than the least stringent allele frequency threshold of 2% used by the 100kGP germline variant prioritisation pipeline.

Ancestry differences in the prioritisation of somatic mutations in actionable genes We compared actionable somatic mutations in 14,532 tumour samples from 13,645 patients. 84% of patients with European ancestry had a somatic mutation in an actionable gene compared with 74% in African, 80% in South Asian and 86% in East Asian ancestry groups (Table 1). We found evidence that fewer mutations in actionable genes were identified for patients of non-European ancestry compared to Europeans when adjusting for sex and cancer type (LRT p=0.007, rate ratio=0.87 for African and 0.80 for South Asian compared to European ancestry; Supplementary Figure 4A).

**Table 1.**
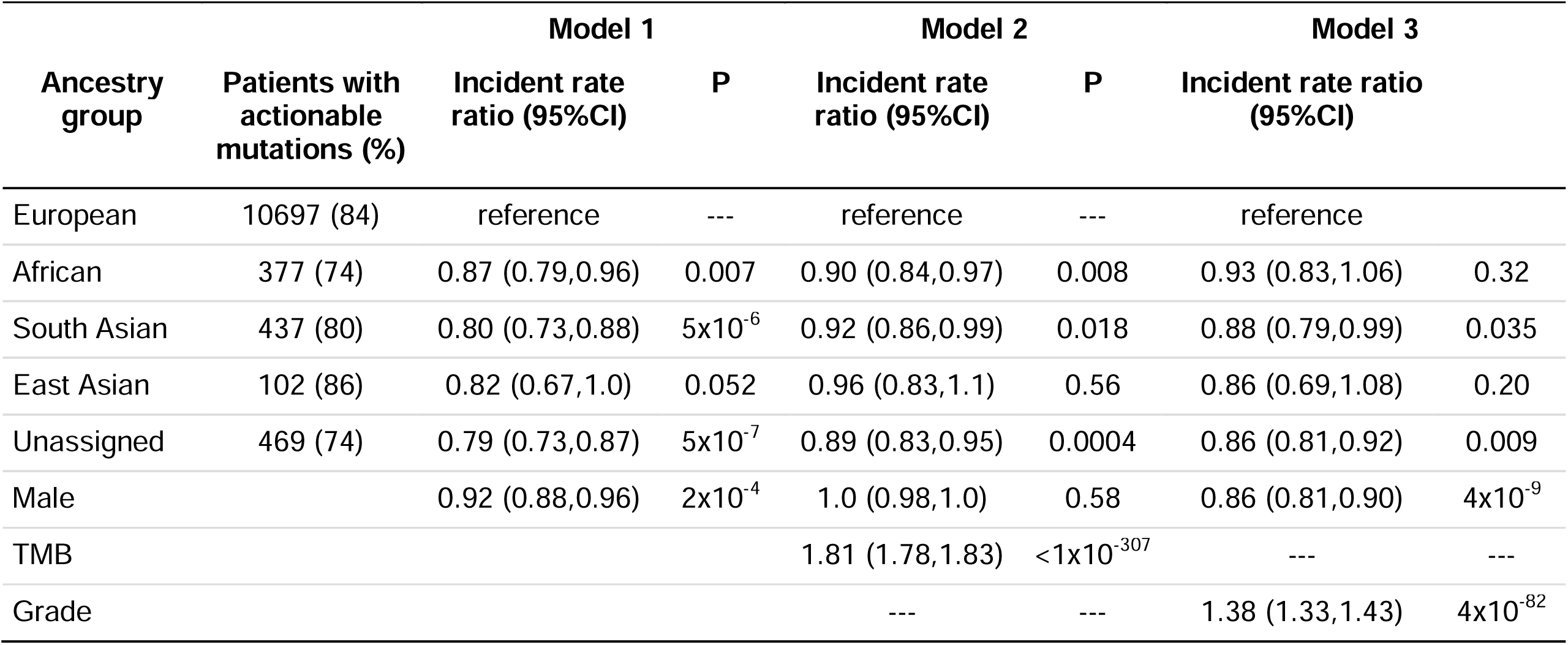
Association between genetically inferred ancestry group and the number of total actionable somatic mutations identified in tumour tissue. First negative binomial regression model adjusted for sex and cancer type (N=14,532 somatic samples from 13,645 patients). Second model adjusted for sex, cancer type and tumour mutation burden (TMB). Model three adjusted for sex, cancer type and tumour grade. European ancestry and female sex were used as references. Note that model 3 was affected by missing data, with a 40% reduction in sample size compared with model 1 (see Supplementary Table 11).

| Ancestry group | Patients with actionable mutations (%) | Model 1 |  | Model 2 |  | Model 3 |  |
| --- | --- | --- | --- | --- | --- | --- | --- |
|  |  | Incident rate ratio (95%CI) | P | Incident rate ratio (95%CI) | P | Incident rate ratio (95%CI) |  |
| European | 10697 (84) | reference | --- | reference | --- | reference |  |
| African | 377 (74) | 0.87 (0.79,0.96) | 0.007 | 0.90 (0.84,0.97) | 0.008 | 0.93 (0.83,1.06) | 0.32 |
| South Asian | 437 (80) | 0.80 (0.73,0.88) | $5 \times 10^{-6}$ | 0.92 (0.86,0.99) | 0.018 | 0.88 (0.79,0.99) | 0.035 |
| East Asian | 102 (86) | 0.82 (0.67,1.0) | 0.052 | 0.96 (0.83,1.1) | 0.56 | 0.86 (0.69,1.08) | 0.20 |
| Unassigned | 469 (74) | 0.79 (0.73,0.87) | $5 \times 10^{-7}$ | 0.89 (0.83,0.95) | 0.0004 | 0.86 (0.81,0.92) | 0.009 |
| Male | | 0.92 (0.88,0.96) | $2 \times 10^{-4}$ | 1.0 (0.98,1.0) | 0.58 | 0.86 (0.81,0.90) | $4 \times 10^{-9}$ |
| TMB | | | | 1.81 (1.78,1.83) | $< 1 \times 10^{-307}$ | --- | --- |
| Grade | | | | --- | --- | 1.38 (1.33,1.43) | $4 \times 10^{-82}$ |

We hypothesized that the reduced numbers of actionable somatic mutations in non-European ancestries were at least partially due to ancestry differences in tumour characteristics. Whilst tumour stage was not associated with the number of somatic mutations in actionable genes (p=0.81), higher tumour grade was (Tables 1; p=4×10^-82^). For some cancer types, there were ancestry differences in terms of tumour grade (Supplementary Figure 5, Supplementary Table 9), with sarcoma tumour grade significantly lower in individuals of South Asian ancestry (p=1×10^-5^), and breast cancer tumour grade higher in women of African ancestry (p=0.016), both compared to European ancestry. Ancestry still had a significant effect on the number of actionable mutations after accounting for tumour grade (LRT p=0.013, Table 1). However, grade was only available for ∼60% of tumour samples (9,278 samples across 8,845 patients) resulting in a significant loss of statistical power, and grade data were more likely to be missing for patients of African and unassigned ancestry and younger patients (Supplementary Table 10).

Therefore, we also considered TMB which was available for the full sample. TMB was associated with an increased number of actionable somatic mutations (p<1×10^-307^; Table 1). However, ancestry remained a significant predictor in the model adjusting for TMB (LRT p=6×10^-4^), with the African and South Asian ancestry groups having significantly fewer actionable mutations (Table 1). In addition, older age was associated with the number of actionable somatic mutations. However, the effect of ancestry was still significant in models adjusted for age (LRT p=0.031).

Findings for somatic mutations in any genes (domains 1, 2 and 3 combined) were consistent with those for mutations in actionable genes (Supplementary Figure 4B, Supplementary Table 11).

## Discussion

This is the first study assessing differences in variant prioritisation related to genetic ancestry in clinical whole-genome sequencing for a national cancer cohort. Given its role as the pilot for nation-wide implementation of clinical whole-genome sequencing in cancer care, the representativeness of the 100,000 Genomes Project is of great importance. Other large studies assessing ancestry differences in cancer sequencing have suffered from underrepresentation of minority ethnic groups^27^. In the 100kGP recruitment rates for Black and Asian ethnicities compared with White ethnicity were consistent with rates in England, except for bladder and prostate (Black and Asian) and breast (Asian only) where Black and Asian ethnicities were recruited at higher rates than expected compared to White ethnicity. This suggests an over-recruitment of minority ethnicities relative to national incidence rates for these cancers. We also found higher average ages in patients with European ancestry which is consistent with ONS statistics demonstrating demographic changes in England over time due to immigration and mixing^28^. The 100kGP may be relatively representative of the general cancer population due to its size and design where participation was not conditional on patient characteristics. However, as a limitation of our study, we cannot rule out the possibility that there are some recruitment biases affecting numbers to a smaller extent, in particular for less-common cancers.

We found evidence that more germline variants in cancer susceptibility genes were prioritised for individuals of non-European ancestry. This was not explained by ancestry differences in genomic diversity or segregation mode. There is little prior evidence that cancer susceptibility variants are generally more frequent in non-European groups except for specific founder effects in some groups ^29^. Therefore, it is more likely that the excess of prioritised variants represents false positives. This implies higher numbers of variants of uncertain significance (VUS) in the non-European ancestry groups, in line with other studies. For example, using data from 5026 patients from SEER in the US, Kurian et al found VUS for 23.7% of White patients but 44.5 % of Black and 50.9% of Asian patients^9^. An excess of prioritised variants may have clinical disadvantages for patients with non-European ancestry since they may make it more difficult to identify true PVs.

A recent US-based study using data from 15,755 patients found lower rates of PVs in Asian and Black patients (12.5% and 12.4% respectively) compared with White patients (15.5%)^10^. They included 76 genes from the MSK-IMPACT panel. Unlike our study, these analyses did not adjust for covariates. We hypothesise that the most likely explanation for the opposite direction of effect in this study are the different strategies used for variant prioritisation. In the MSK study, “variants were independently assessed and manually curated to define” PVs. Given the underrepresentation of non-European ancestry groups in genetic research, this may favour variants that are sufficiently frequent in European ancestry groups whilst variants relevant to other groups may be understudied and therefore not included in the curated list. This bias is less likely to affect our cohort which includes both predicted loss-of-function mutations and those annotated in ClinVar as pathogenic or likely pathogenic with filtering out of variants classified as benign or likely benign.

We found that up to 25% of prioritised candidate variants in the non-European ancestry groups were common based on ancestry-specific reference data. These are unlikely to be pathogenic. Future strategies to reduce the excess of prioritised variants in these groups could be to separate reference data sets into ancestry groups for filtering out common variants. Furthermore, it is important to continue to increase sample sizes and diversity of reference resources.

On the other hand, we found that fewer somatic mutations in potentially actionable genes were prioritised by the cancer bioinformatics pipeline in individuals of non-European ancestry. These differences remained significant when accounting for cancer type, sex and age. This is in line with findings from studies in the US^27^. Using data from 45,000 pan-cancer patients from the MSK cohort, Arora et al found actionable mutations in 30% of African vs 33% of European ancestry patients^27^. The differences were reduced but remained significant when the authors accounted for cancer subtype. Some genes, such as *EGFR*, show differential rates of somatic mutations in tumours across ancestry groups^30, 31^. It is possible that the underrepresentation of diverse ancestry groups in research studies has favoured discovery of variants in actionable genes that are more frequent in European ancestry groups.

There are also ethnicity differences for some tumour characteristics. For example Black women in the UK present more frequently with triple negative breast cancer^32^. When accounting for tumour grade or TMB, the differences between European and other ancestry groups reduced but the number of mutations in actionable genes remained consistently lower in African and South Asian ancestries. This suggests that the different detection rates of actionable mutations may be partially mediated by differences in tumour characteristics.

However, as a limitation of our study, missing data made it difficult to assess the mediating role of tumour features more widely. For example, grade was only available for ∼60% of tumour samples and this missingness might not have been entirely random, as patients of African ancestry were more likely to have missing grade.

Further study limitations include the arbitrary nature of assigning individuals to ancestry groups based on genetic similarity to reference groups. Furthermore, the intersection of ancestry and cancer types led to small group sizes for some combinations. Therefore, most of the analyses adjusted for cancer type but did not consider them separately. It is likely that not all the conclusions apply equally across all cancer types.

In conclusion, we found that ethnicity in the 100kGP cancer programme is largely representative of England, distinguishing it as a uniquely useful pan-cancer cohort for research on the role of ethnicity in diagnosis, prognosis and care in the wider cancer population. We also identified statistically significant ancestry differences in the detection of both germline PVs and somatic mutations in actionable genes. Opportunities for further research will increase with the release of additional data from routine sequencing as part of the NHS Genomic Medicine Service. Our study underlines the need for this, if we are to ensure equitable benefits from genomics in cancer care for diverse populations.

## Acknowledgements

This research was made possible through access to data in the National Genomic Research Library, which is managed by Genomics England Limited (a wholly owned company of the Department of Health and Social Care). The National Genomic Research Library holds data provided by patients and collected by the NHS as part of their care and data collected as part of their participation in research. The National Genomic Research Library is funded by the National Institute for Health Research and NHS England. The Wellcome Trust, Cancer Research UK and the Medical Research Council have also funded research infrastructure. KK was also supported by the European Union under the Horizon 2020 research and innovation programme (No 948561).

## References

1. NHS Genomic Medicine Service [Internet][cited 2024 Feb 6] Available from: https://www.england.nhs.uk/genomics/nhs-genomic-med-service/

2. Turnbull C, Scott RH, Thomas E, et al: The 100 000 Genomes Project: bringing whole genome sequencing to the NHS [Internet]. BMJ 361, 2018Available from: https://www.bmj.com/content/361/bmj.k1687

3. Sosinsky A, Ambrose J, Cross W, et al: Insights for precision oncology from the integration of genomic and clinical data of 13,880 tumors from the 100,000 Genomes Cancer Programme. Nat Med 30:279–289, 2024

4. Alarcón Garavito GA, Moniz T, Déom N, et al: The implementation of large-scale genomic screening or diagnostic programmes: A rapid evidence review. Eur J Hum Genet 31:282–295, 2023

5. Delon C, Brown KF, Payne NWS, et al: Differences in cancer incidence by broad ethnic group in England, 2013–2017. Br J Cancer 126:1765–1773, 2022

6. DeSantis CE, Miller KD, Goding Sauer A, et al: Cancer statistics for African Americans, 2019. CA Cancer J Clin 69:211–233, 2019

7. Aizer AA, Wilhite TJ, Chen M-H, et al: Lack of reduction in racial disparities in cancer-specific mortality over a 20-year period. Cancer 120:1532–1539, 2014

8. Anna Fry, Becky White, Diana Nagarwalla, et al: Relationship between ethnicity and stage at diagnosis in England: a national analysis of six cancer sites. BMJ Open 13:e062079, 2023

9. Kurian AW, Ward KC, Hamilton AS, et al: Uptake, Results, and Outcomes of Germline Multiple-Gene Sequencing After Diagnosis of Breast Cancer. JAMA Oncol 4:1066–1072, 2018

10. Liu YL, Maio A, Kemel Y, et al: Disparities in cancer genetics care by race/ethnicity among pan-cancer patients with pathogenic germline variants. Cancer 128:3870–3879, 2022

11. Domchek SM, Yao S, Chen F, et al: Comparison of the Prevalence of Pathogenic Variants in Cancer Susceptibility Genes in Black Women and Non-Hispanic White Women With Breast Cancer in the United States. JAMA Oncol 7:1045–1050, 2021

12. Genomics England: The National Genomics Research Library

13. gov.uk: List of ethnic groups [Internet][cited 2023 Nov 6] Available from: https://www.ethnicity-facts-figures.service.gov.uk/style-guide/ethnic-groups

14. Office for National Statistics: 2011 Census analysis: Ethnicity and religion of the non-UK born population in England and Wales: 2011 [Internet], 2015[cited 2023 Nov 6] Available from: https://www.ons.gov.uk/peoplepopulationandcommunity/culturalidentity/ethnicity/articles/2011censusanalysisethnicityandreligionofthenonukbornpopulationinenglandandw ales/2015-06-18

15. Office for National Statistics: Population denominators by ethnic group, regions and countries: England and Wales, 2011 to 2018 [Internet], 2018[cited 2023 Nov 6] Available from: https://www.ons.gov.uk/peoplepopulationandcommunity/populationandmigration/populationestimates/adhocs/008780populationdenominatorsbyethnicgroupregionsandcountriesenglandandwales2011to2017

16. Pace M, Lanzieri G, Glickman M, et al: Revision of the European standard population. Rep Eurostat Task ForcePublications Off Eur Union 2013 20, 2013

17. Altman DG, Bland JM: Interaction revisited: the difference between two estimates. Bmj 326:219, 2003

18. Kousathanas A, Pairo-Castineira E, Rawlik K, et al: Whole-genome sequencing reveals host factors underlying critical COVID-19. Nature 607:97–103, 2022

19. Ellen McDonagh et al.: Cancer Analysis Technical Information Document v1.11 [Internet], 2019 Available from: https://files.genomicsengland.co.uk/forms/Cancer-Analysis-Technical-Information-Document-v1-11-main.pdf

20. Genomics England: AggV2 functional annotation [Internet][cited 2024 Apr 6] Available from: https://re-docs.genomicsengland.co.uk/functional_annotation

21. Martin AR, Williams E, Foulger RE, et al: PanelApp crowdsources expert knowledge to establish consensus diagnostic gene panels. Nat Genet 51:1560– 1565, 2019

22. Landrum MJ, Lee JM, Benson M, et al: ClinVar: improving access to variant interpretations and supporting evidence. Nucleic Acids Res 46:D1062–D1067, 2018

23. Brooks ME, Kristensen K, Benthem KJ van, et al: glmmTMB Balances Speed and Flexibility Among Packages for Zero-inflated Generalized Linear Mixed Modeling. R J 9:378–400, 2017

24. Karczewski KJ, Francioli LC, Tiao G, et al: The mutational constraint spectrum quantified from variation in 141,456 humans. Nature 581:434–443, 2020

25. Wall JD, Sathirapongsasuti JF, Gupta R, et al: South Asian medical cohorts reveal strong founder effects and high rates of homozygosity. Nat Commun 14:3377, 2023

26. Peto J, Collins N, Barfoot R, et al: Prevalence of BRCA1 and BRCA2 gene mutations in patients with early-onset breast cancer. J Natl Cancer Inst 91:943–949, 1999

27. Arora K, Tran TN, Kemel Y, et al: Genetic Ancestry Correlates with Somatic Differences in a Real-World Clinical Cancer Sequencing Cohort. Cancer Discov 12:2552–2565, 2022

28. Office for National Statistics: Population of England and Wales [Internet]. Popul Engl Wales, 2022[cited 2023 Oct 27] Available from: https://www.ethnicity-facts-figures.service.gov.uk/uk-population-by-ethnicity/national-and-regional-populations/population-of-england-and-wales/latest

29. Neuhausen SL: Ethnic differences in cancer risk resulting from genetic variation. Cancer Interdiscip Int J Am Cancer Soc 86:2575–2582, 1999

30. Shigematsu H, Lin L, Takahashi T, et al: Clinical and Biological Features Associated With Epidermal Growth Factor Receptor Gene Mutations in Lung Cancers. JNCI J Natl Cancer Inst 97:339–346, 2005

31. Carrot-Zhang J, Soca-Chafre G, Patterson N, et al: Genetic Ancestry Contributes to Somatic Mutations in Lung Cancers from Admixed Latin American Populations. Cancer Discov 11:591–598, 2021

32. Gathani T, Reeves G, Broggio J, et al: Ethnicity and the tumour characteristics of invasive breast cancer in over 116,500 women in England. Br J Cancer 125:611– 617, 2021

